# WBC population dynamics differ in response to acute ischemic and infectious insults and can discriminate clinically maladaptive responses to myocardial infarction

**DOI:** 10.64898/2026.05.03.26352288

**Authors:** Sylvia Ranjeva, Brody Foy, Christopher Mow, Jonathan Carlson, Aaron Aguirre, Matthias Nahrendorf, John Higgins

## Abstract

Leukocytosis, or elevated white blood cell count (WBC), is a clinical hallmark of the systemic inflammatory response following acute ischemia or infection. However, WBC population dynamics during the initial inflammatory response are poorly understood. It is unknown whether early WBC dynamics differ by etiology or impact clinical risk. We fit mathematical models to WBC trajectories for patients hospitalized following acute ischemia or infection. We found differences between responses to strong ischemic insult (acute myocardial infarction, AMI) and strong infectious insult (sepsis). Among patients who recovered and survived hospitalization, net WBC growth following ischemia was ∼1.8x faster than following infection. Response kinetics and dynamics were correlated with short-term mortality in AMI. Increased immature neutrophil production over 24h preceding WBC peak was associated with ∼2x increased odds of short-term AMI mortality, suggesting that dysregulated responses to ischemia may involve bone marrow overactivation towards increased proliferation. Our work provides novel insight into fundamental etiology-specific differences in acute inflammatory responses from routine clinical data and is consistent with recent evidence of a maladaptive role for emergency granulopoiesis in AMI.

**One Sentence Summary:** WBC dynamics differ between ischemia and infection in the early response to acute inflammatory insult, and can discriminate clinically maladaptive responses to myocardial infarction.

## INTRODUCTION

Acute inflammatory responses are central to the pathophysiology of ischemic and infectious insults. In clinical settings, elevated white blood cell (WBC) count, or leukocytosis, is a key non-specific signal of systemic inflammation. However, the dynamics of early WBC responses to inflammatory insults are poorly understood(*1*). While clinicians use WBC changes as a sign of inflammation, it is generally not possible to distinguish WBC trajectories among possible causes, like ischemia or infection, and it is unclear how these response-phase dynamics may influence clinical risk. Excess neutrophil activity is implicated in both strong infectious insult (e.g. sepsis(*2*)) and strong ischemic insult (e.g. acute myocardial infarction, AMI(*3*)), motivating current investigations into immunomodulatory therapies. Static clinical markers of excess neutrophil activity (e.g neutrophil to lymphocyte ratio, NLR) are prognostic in sepsis, AMI, and stroke(*4, 5*). However, it is unclear whether early dynamics of these WBC subpopulations differ between inflammatory etiologies or influence clinical risk. Overall, etiology-specific WBC trajectories may reflect key differences in WBC production, demargination, and clearance. Understanding these differences may reveal important cell population-level immunologic mechanisms and enhance early risk-stratification.

A recent analysis from our group showed that patients who mount an effective acute inflammatory response to infection, ischemia, or trauma follow a universal recovery trajectory after reaching a maximum WBC count(*6*), characterized by correlated dynamics of WBC and platelet populations, with approximately exponential WBC decay and linear platelet count growth after a ∼1-2d delay. The consistency of WBC trajectories during recovery from acute inflammation contrasts with heterogeneity during the earlier phase of inflammatory responses.(*5*) This observed heterogeneity(*7, 8*) implies important etiology-specific differences in the early hematologic dynamics preceding WBC peak, before the recovery trajectory is initiated.

Here, we leverage mathematical models of WBC kinetics to investigate the early inflammatory response among patients hospitalized following acute ischemic or infectious insults and characterize etiology-specific dynamics of WBC subpopulations that correlate with clinical risk. As WBCs are the primary effectors of inflammation, we focus on their population dynamics in these settings, prior to initiation of inflammatory recovery. We take advantage of our recent characterization of hematologic recovery trajectories (*6*) to focus on the early WBC response preceding WBC peak.

## RESULTS

### Data and statistical models

We analyzed clinical laboratory data from adults admitted to Massachusetts General Hospital (1/2016 -12/2020) with one of four infectious or ischemic disorders (sepsis, non-fulminant *C. difficile* colitis, ischemic stroke, or AMI) (Methods), focusing on the previously defined cohorts that were used in our prior analysis of hematologic recovery dynamics(*6*). 2554 patients (*n=*584 sepsis, *n=*161 C. difficile, *n=*703 stroke, *n=*1106 AMI) met inclusion criteria. WBC response trajectories (Fig 1, A and B) were well-represented by exponential growth (Fig 1, C and D), which provided superior fits compared to other simple models (Supplemental Material, section S1, Fig. S1). Models were fit to individually scaled WBC counts to account for differences in patient baseline levels (Methods, Supplemental Material, Fig. S2), consistent with our prior analyses(*6,29*). Among the AMI cohort, median time between the ischemic event (corresponding to peak troponin) and peak WBC was three days (Supplemental Material, Fig. S3).

**Fig. 1.**
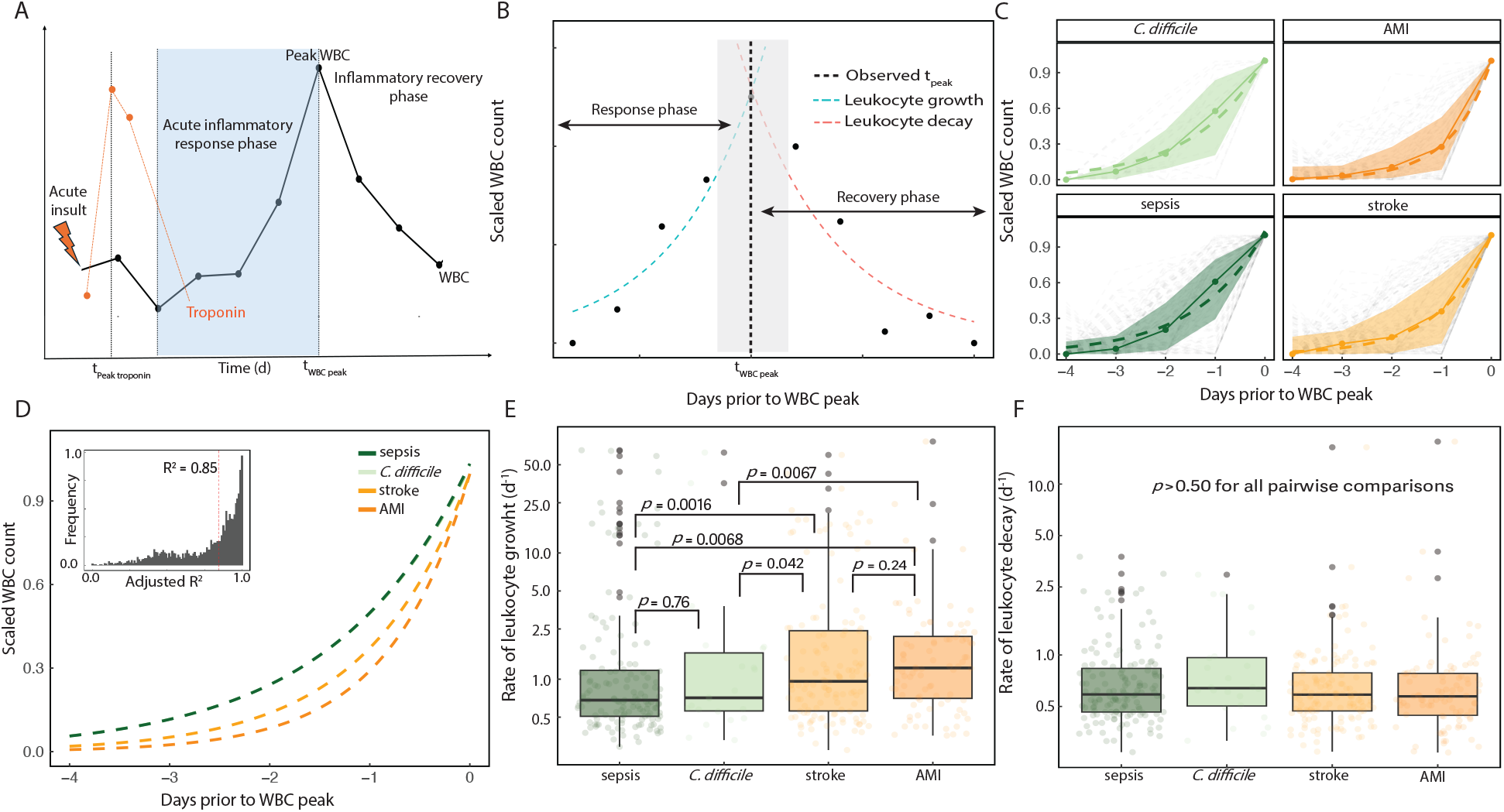
WBC growth rate is faster in healthy response to ischemia compared to healthy response to infection. **A**. Schematic of inflammatory response for one individual with acute myocardial infarction (AMI). After the ischemic event, troponin rapidly peaks, followed by an acute WBC response. Peak WBC marks the beginning of recovery, and WBC returns to baseline. **B**. Schematic for WBC response and recovery for a representative individual with sepsis, with exponential fits for WBC growth and decay. Gray shaded area denotes uncertainty around the true WBC peak due to sampling frequency. **C**. WBC response trajectories among the favorable trajectory cohort (*n*=382) for patients with infectious (22 *C. diff* colitis and 139 sepsis) or ischemic (88 AMI and 133 stroke) conditions. Solid lines show the median across individuals at each timepoint, dashed lines show an aggregate exponential fit, and shaded regions denote the interquartile range (IQR). Light grey dashed lines show individual trajectories. **D**. Aggregate exponential fit of etiology-specific WBC response trajectories. Inset shows a histogram of adjusted coefficients of determination (R^2^) for exponential WBC growth models. For 62% of individuals, the exponential model explains >85% of the variance in the response-phase WBC measurements. **E**. Boxplots of inferred exponential WBC growth rates (log scale). Black horizontal lines denote the median. Shaded areas give the IQR. **F**. Boxplots of fitted exponential WBC decay rates (log scale) during the recovery phase.

### Etiology-specific differences in early WBC dynamics among favorable clinical responses

To investigate baseline differences between responses to infection and ischemia, we first assessed ‘favorable’ WBC responses defined as those from the subgroup of patients (*n=*382) with well-fitting hematologic growth and recovery trajectories (Methods) who survived to 30d. See Supplemental Material, Section S3, for example clinical courses for patients identified to have favorable and unfavorable trajectories. Among this favorable trajectory cohort, WBC growth rate varied across etiologies (Fig 1 C-E, Supplemental Material, Section S2, Fig. S4), from lowest in sepsis (strong infectious insult) to highest in AMI (strong ischemic insult). WBC recovery rates were indistinguishable across etiologies (Fig. 1F), and inferred WBC growth rates were not associated with WBC decay rates (Supplemental Material, Fig. S5).

### Etiology-specific differences in early WBC dynamics among maladaptive clinical responses

The faster WBC response to ischemia among the favorable trajectory cohort suggested that rapid WBC population growth, net of clearance, may be more important for effective immune responses to ischemic insult than for responses to infectious processes. Having established etiology-specific differences in pre-WBC peak leukocyte dynamics among clinically favorable trajectories, we next investigated clinically maladaptive trajectories, focusing on strong ischemic insult (AMI) and strong infectious insult (sepsis). We analyzed WBC response trajectories stratified according to 30d mortality (see Methods). There were 100 non-survivors (9%) in the AMI group and 77 (13%) in the sepsis group. For AMI, but not for sepsis, a faster rate of early WBC response was significantly associated with increased 30d survival (Fig. 2A). Furthermore, the lower WBC growth rate in non-surviving AMI patients was similar to that for the sepsis population (Fig. 2A).

**Fig. 2.**
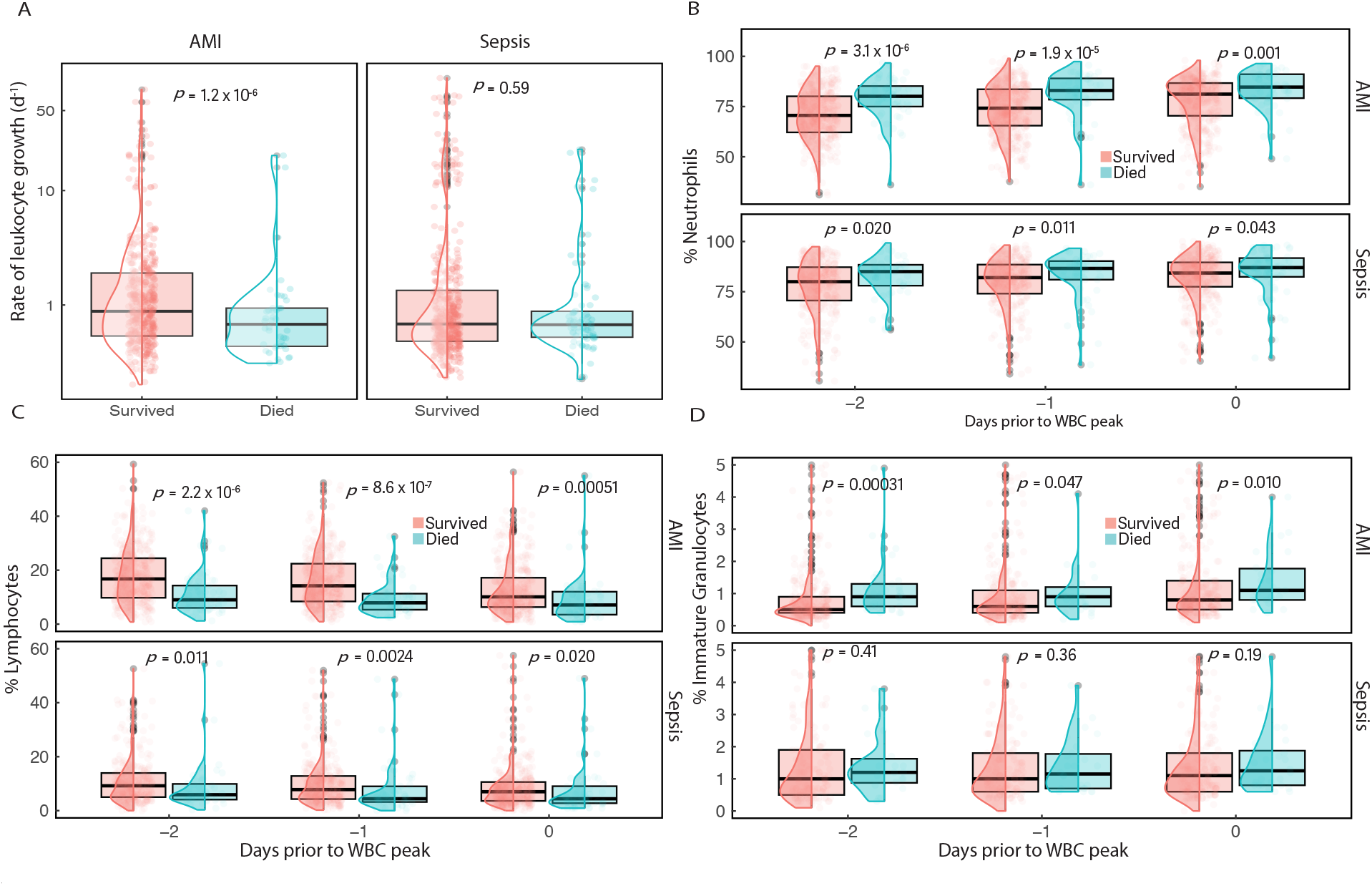
Faster WBC growth rate is associated with survival in AMI, but not sepsis, and differences in *de novo* neutrophil production correlate with AMI survival. **A**. Boxplots and overlaid distributions show that exponential WBC growth rates in AMI (left) are higher in surviving patients (red, *n =* 1006) compared to non-survivors (cyan, *n* = 100), but WBC growth rates in sepsis (right) are not associated with 30d survival. Values shown on log scale. Black horizontal lines give the median across patients. Shaded boxes give 95% confidence intervals. Panels **B-D** give boxplots and overlaid distributions showing neutrophil fractions (B), lymphocyte fractions (C), and immature granulocyte fractions (D) over three days prior to WBC peak.

### Excess immature neutrophil activity and short-term mortality following acute myocardial infarction

Given that slower WBC population growth was associated with mortality in AMI, but not sepsis, we explored whether this difference could be attributed to specific WBC lineages. In both AMI and sepsis, mortality was associated with neutrophilic predominance and relative lymphopenia close to WBC peak (Fig. 2, B-C, Methods). For both AMI and sepsis, patients with higher percent neutrophils were less likely to survive. Among surviving patients, neutrophil fractions and immature granulocyte fractions (IGF, an estimate of *de novo* neutrophil production(*9*)) close to WBC peak were consistently lower in AMI compared to sepsis (Supplemental Material, Fig. S6 and S7), while among non-surviving patients, neutrophil and IGF were indistinguishable between AMI and sepsis. These trends held for analyses using absolute counts of WBC sub-populations (Supplemental Material, Fig. S8). For AMI, survivors experienced a significantly greater absolute increase in circulating neutrophils over the 24h prior to WBC peak compared to non-survivors (Supplemental Material, Fig. S9). In the sepsis population, there was no mortality-specific difference in neutrophil population growth close to WBC peak (Supplemental Material, Fig. S9). Overall, the association of faster increase in circulating neutrophil populations with AMI survival suggests that neutrophil dynamics are likely a major contributor to the faster net WBC growth among AMI survivors.

Neutrophil populations can increase as a result of *de novo* production or demargination of pre-existing neutrophil pools or as a result of decreases in baseline cell turnover or margination (*10*). To explore the potential contribution of *de novo* neutrophil production to rapid neutrophil population growth in AMI survivors, we analyzed measurements of IGF(*9*). Prior evidence suggests that demargination of pre-existing neutrophils as opposed to *de novo* production is critical in the effective early response to ischemic injury(*11, 12*). Consistent with this evidence, AMI survivors had significantly lower IGF over all three days prior to WBC peak than non-survivors (Fig. 2D). These results, while correlative, are consistent with the idea that rapid demargination of neutrophils is important for healthy responses to ischemia, while increased *de novo* production of neutrophils, a slower process, may be associated with a maladaptive response to ischemia.

Since increased IGF preceding WBC peak appeared to be associated with maladaptive AMI responses, we investigated associations between IGF and mortality in the AMI cohort (Fig. 3).

**Fig. 3.**
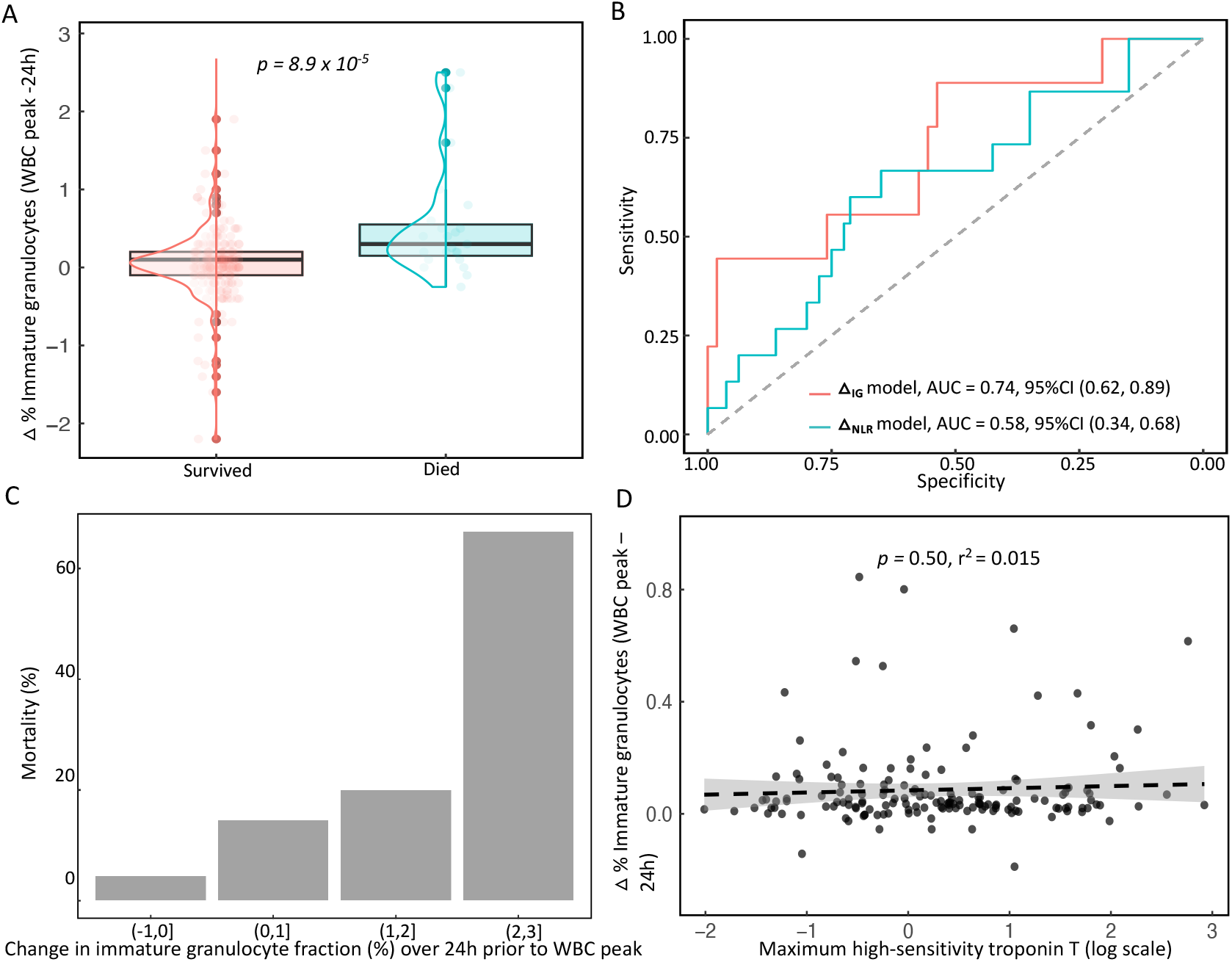
The magnitude of change in the immature granulocyte fraction (IGF) is associated with 30d survival in AMI. **A**. Boxplots and distributions show changes (Δ_IG_) in the immature granulocyte fraction (IGF) during the 24h prior to WBC peak among patients with AMI. **B**. Receiver operating characteristic (ROC) curves show performance of Δ_IG_ as a predictor of 30d mortality compared to performance of the neutrophil: lymphocyte ratio (Δ_NLR_ model). Area under the curve (AUC) is calculated along with its 95% confidence interval, constructed by bootstrapping (10,000 replicates). **C**. Short term mortality exhibits a dose-response correlation with increasing Δ_IG_ over the 24h prior to WBC peak. **D**. Δ_IG_ was not significantly correlated with peak high-sensitivity troponin T among patients (*n* = 243) that had consistent consecutive cell count differentials prior to WBC peak and an observed rise to peak in high-sensitivity troponin T during admission. Black dashed line gives the fitted linear regression. Shaded gray area denotes the standard error.

Greater increase in IGF over 24h pre-WBC peak (Δ_IG_) was associated with short-term AMI mortality (Fig. 3A), including in an age-adjusted multivariate logistic regression model (OR 2.04, 95% CI [1.41, 3.13]). Δ_IG_ showed stronger association with mortality than the neutrophil:lymphocyte ratio (NLR) and similar association to that for the recently reported Platelet:WBC ratio(*13*) (Fig. 3B and Supplemental Material, Section S5, Fig. S10). Furthermore, short-term mortality risk scaled with Δ_IG_ in a dose-response-like relationship (Fig. 3C). Because infarct size is known to be associated with mortality(*14*), we investigated whether Δ_IG_ was simply a surrogate for infarct size by comparing it to an established correlate of infarct size, peak troponin(*15*). We found no association (*p*=0.50, Fig. 3D), suggesting that the observed trends in Δ_IG_ provide risk information complementary to the magnitude of initial injury. Consistent with this idea, combining peak troponin with Δ_IG_ increased the accuracy of short-term mortality risk prediction (Supplemental Material, Fig. S11).

### Single-cell signatures of neutrophil immaturity and short-term mortality following acute myocardial infarction

Neutrophil population dynamics close to WBC peak appeared to distinguish effective inflammatory responses from maladaptive responses to AMI. To investigate changes in neutrophil dynamics that may not be reflected in population size, we analyzed single-cell raw data available from CBCs (*16*) measured close to WBC peak (Methods). We studied the multidimensional single-cell distributions of neutrophil populations over time and found that the single-neutrophil “side fluorescence intensity” (NE-SFL), a correlate of nuclear morphology and an index of immaturity, was higher on average in AMI non-survivors than in survivors. CBCs from AMI non-survivors also had a greater variance in NE-SFL (quantified by the NE-WY parameter). NE-WY was greater in AMI non-survivors on each of the three days prior to WBC peak (Supplemental Material, Fig. S12). This pattern is consistent with previous reports finding that these markers of average neutrophil immaturity and its variance differentiated hospitalized patients with septic shock from those with benign infection(*17*) and suggests that robust markers of adaptative and maladaptive inflammatory responses, independent from WBC count, may be present in raw data from routine CBCs.

## DISCUSSION

Inflammation is the fundamental response to cellular damage, and effective inflammatory responses must respond appropriately to diverse etiology, halting further injury, repairing damage, and recovering to baseline promptly and without long-term sequelae. Here, we have identified previously unappreciated differences in early WBC dynamics between inflammatory responses to ischemia and infection. Among clinically favorable trajectories, ischemic conditions induced more rapid and shorter duration WBC population growth, possibly reflecting relatively more demargination of populations of pre-formed leukocytes. Conversely, infectious conditions showed slower WBC growth and greater neutrophilic predominance, possibly reflecting greater bone marrow activity towards de novo neutrophil production. In AMI, deviation from a favorable faster WBC response trajectory was associated with higher short-term mortality.

On a pathophysiologic level, this study is consistent with a hypothesized maladaptive role in AMI for excess immature neutrophil production (18–21). Decades of in vitro and in vivo work have established a complex and delicate balance of neutrophil production, differentiation, and demargination in response to myocardial injury(18–21). Recent evidence supports a pathophysiologic role for emergency hematopoiesis after AMI (20), a bone marrow response characterized by accelerated differentiation of hematopoietic stem cells (HSCs) to facilitate healing. Excess signaling of neutrophil activity and WBC overproduction leads to accelerated cardiomyocyte death and pathologic cardiac remodeling(18). In turn, dysregulation of neutrophil apoptosis at the site of ischemic injury prevents macrophage polarization towards a resolving phenotype(21). Consistent with this paradigm, our work suggests that maladaptive AMI responses may involve enhanced de novo production of immature, undifferentiated neutrophils.

We also find slower growth of the overall WBC population close to the WBC peak, implying less demargination of pre-existing neutrophil pools or increased turnover. This retrospective observational analysis cannot establish a causal role for neutrophil-mediated injury in myocardial remodeling, but the WBC population dynamics in this study cohort are consistent with a state of bone marrow hyper-activation that has previously been characterized in maladaptive AMI responses (19–22). Importantly, the IGF marker of immature granulocyte activity was not associated with peak troponin, suggesting that the observed patterns are not merely a reflection of a larger inflammatory insult.

The post-reperfusion inflammasome is a major focus of immunologic AMI therapies. While broad immunosuppressive agents have shown equivocal effect in randomized controlled trials (23, 24), targeted blockade of cytokines (e.g. IL-1, IL1-ß) that stimulate neutrophil production has shown promise in preventing adverse AMI outcomes including short-term mortality (25–27). Furthermore, blockage of excess emergency hematopoiesis prevents adverse cardiac remodeling in a murine model of AMI (28), highlighting modulation of HSC differentiation as a potential avenue of therapeutic development. Markers of immature neutrophil production may therefore provide complementary benchmarks to guide the expanding use of targeted immunomodulatory therapies.

Our work has several imitations. Future work with higher-frequency measurements is needed for more precise characterization of WBC kinetics. This analysis required availability of multiple CBCs, and prospective follow-up studies are needed to increase confidence in the robustness of the identified patterns. In particular, identification of the true WBC peak was limited by the daily observation interval. Additionally, while we excluded patients with known evidence of coexisting infectious and ischemic diagnoses during the observation period, future study is needed to increase confidence that the sepsis-like dynamics associated with poorer prognosis in AMI do not reflect the presence of subclinical infectious agents. We found no significant effect of AMI type on the inferred WBC dynamics in maladaptive trajectories (Supplemental Material, Section S4, Fig. S13), but further study under controlled conditions is warranted, as the observation structure required to model early WBC dynamics restricted our analysis largely to patients with non-ST-elevation MI (NSTEMI). Finally, as above, we cannot make claims of causality due to the observational nature of this study.

Overall, we present novel evidence for fundamental etiology-specific differences in leukocyte population dynamics following acute inflammatory insult. Our work suggests that leukocyte programming towards enhanced immature neutrophil production in AMI, consistent with excessive emergency hematopoiesis, can be detected at the level of cell population dynamics using routine clinical data and is associated with poorer clinical outcomes. Our study motivates more extensive single-WBC analysis from routine and prospective CBCs to elucidate mechanisms responsible for altered neutrophil population dynamics in both effective and maladaptive inflammatory responses.

## MATERIALS AND METHODS

### Patient data and inclusion criteria

This work expands prior analysis that established universal hematologic recovery trajectories among patients facing acute inflammatory insult, drawing from the previously described patient cohort.(*6*)

Briefly, hematologic data was collected from adults admitted to Massachusetts General Hospital between 1/1/2016–12/31/2020, with the current analysis focusing on patients with one of four infectious or ischemic disorders: sepsis, non-fulminant *C. difficile* colitis, ischemic stroke, or acute myocardial infarction (AMI). Admission-associated daily complete blood counts (CBC) and differential blood counts for each cohort were collected from the Research Patient Data Registry and Enterprise Data Warehouse, along with demographics, admission and discharge dates, and all-cause mortality within 30 days of hospital discharge. Key inclusion criteria were age >18y and hospital length of stay >48h. Diagnoses were confirmed via ICD codes (ICD-9 and ICD-10). Stroke was defined as any diagnosis of a stroke or cerebrovascular accident. The colitis cohort was limited to patients with a diagnosis of C. difficile colitis or infectious colitis, or with a diagnosis of colitis in conjunction with a positive C. difficile toxin assay. The sepsis cohort included any patients with clinical diagnosis of sepsis regardless of infectious organism. The myocardial infarction cohort included patients with an admitting diagnosis of ST segment elevation or non-ST segment elevation myocardial infarction. Patients with overlapping concurrent diagnoses during the observation period were excluded. Among the AMI cohort, 910 patients had available high sensitivity troponin-T data in the EMR during admission (for 196 AMI patients that met inclusion criteria, hs-troponin was not available with routine CBC data, for example if collected at an outside emergency department prior to transfer for admission to this tertiary care center).

Patients were temporally aligned according to peak white blood cell (WBC) count within 14 days of admission. For the results requiring modeling of both the hematologic early inflammatory response and recovery trajectory, we restricted analysis to patients with >3 days of CBC data prior to WBC peak and >5 days of CBC data post-WBC peak. Data for leukocyte sub-populations (neutrophils, lymphocytes, and immature granulocytes fractions) were obtained from clinical CBC differentials measured on Sysmex XN-9000 instruments.

### Models of hematologic response and recovery dynamics

The leukocyte response phase (Fig. 1A) was defined over 4 days prior to WBC peak. The starting point for the response phase was selected empirically based on observed patterns of cohort-level increases in WBC counts (Supplemental Material, Fig. S14). Consistent with the prior analysis of hematologic recovery trajectories(*6*), inflammatory recovery trajectories were modeled according to WBC and lagged PLT dynamics following WBC peak. The inflammatory response phase was modeled with earlier WBC trajectories prior to WBC peak. We focused solely on WBC dynamics during the early inflammatory response, as leukocytes are the primary effectors of the early response. Empirically, platelet activity was widely heterogenous prior to the initiation of the ordered recovery trajectory (Supplemental Material, Fig. S15).

To model WBC dynamics, models of exponential growth and decay:

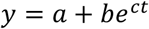

with rate parameter *c* were fitted to individual leukocyte trajectories pre-WBC peak (c>0) and post-WBC peak (c < 0), scaling the absolute WBC to normalize dynamics across patients:

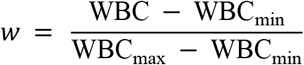

Platelet recovery dynamics were modeled as lagged linear growth following WBC peak

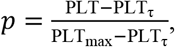

where timepoint τ reflects an inferred time lag for linear platelet recovery. Goodness of model fit was assessed by adjusted R^2^.

### Definition of a ‘favorable trajectory’ cohort

To investigate baseline differences between early inflammatory responses to ischemia and infection, we first defined a cohort of patients with ‘favorable’ hematologic trajectories. We selected patients according to three criteria:

1. Well-fitting hematologic recovery trajectories, indicating ordered resolution of the inflammatory response. Patients were included according to the goodness-of-fit of the WBC and platelet recovery models, requiring adjusted R^2^ > 0.85 for both exponential WBC decay and linear platelet growth.
2. Well-fitting WBC response-phase trajectory, with R^2^ > 0.85 for the exponential WBC growth model.
3. Survival of at least 30d.

A total of 382 patients met all three criteria for the main analysis. See Supplementary Sections S2 and S3 for more detail.

### Statistical analysis

We compared etiology-specific WBC growth rates, stratified by 30-day mortality, and leukocyte sub-populations (comparisons by Wilcoxon rank sum test, α=0.05). Dependence of short-term mortality risk on immature granulocyte activity was assessed via multivariate logistic regression and area under the receiver operating characteristic curve (AUC, using a 30% internal validation cohort, with 95% confidence intervals generated by bootstrapping with 10,000 replicates). All statistical analyses were performed in R (v4.0.2).

### Analysis of single-cell CBC data

Single-cell population data were analyzed from clinically obtained CBC samples for the AMI cohort measured on Sysmex XN-9000 automated hematology instruments (Sysmex Corporation, Kobe, Japan). Briefly, the leukocyte differential channel (WDF channel) measures single-cell optical signals for 1000s of WBCs and uses them to derive the WBC differential. Optical signals are obtained following incubation with surfactant reagents and a fluorescent dye. We analyzed single cell distributions along two axes: side scatter (x-axis) and side fluorescence intensity (y-axis), which separate leukocyte populations according to cellular morphology and nucleic acid content, respectively. Specifically, we compared 4 parameters measured for each CBC: mean neutrophil side scatter intensity (NE-SSC), an index of the dispersion of the side scatter signal across the neutrophil population (NE-WX), mean neutrophil fluorescence intensity (NE-SFL), and an index of the dispersion of neutrophil fluorescence (NE-WY). Population-level distributions of these values were compared from serial CBC samples over the 2 days prior to WBC peak.

## Data Availability

All data produced in the present study are available upon reasonable request to the authors.

## Data availability

De-identified study data are available upon reasonable request from the corresponding authors (S.R. and J.H) and depending on IRB approval. Any request will be reviewed promptly by the corresponding authors and be shared under data transfer agreement from the corresponding author’s institution.

## Author contributions

Conceptualization: SLR, JMH

Methodology: SLR, BF, CW, JMH

Investigation: SLR, BF, CM, JC, AA, MN, JMH

Visualization: SLR

Funding acquisition: JMH

Writing – original draft: SLR, JMH

Writing – review & editing: SLR, BF, CM, JC, AA, MN, JMH

## REFERENCES

1. J. C. Marsh, D. R. Boggs, G. E. Cartwright, M. M. Wintrobe, Neutrophil kinetics in acute infection. J. Clin. Invest. 46, 1943–1953 (1967).

2. X. F. Shen, K. Cao, J. P. Jiang, W. X. Guan, J. F. Du, Neutrophil dysregulation during sepsis: an overview and update. Blackwell Publishing Inc. [Preprint] (2017). 10.1111/jcmm.13112.

3. E. Mezzaroma, S. Toldo, D. Farkas, I. M. Seropian, B. W. Van Tassell, F. N. Salloum, H. R. Kannan, A. C. Menna, N. F. Voelkel, A. Abbate, The inflammasome promotes adverse cardiac remodeling following acute myocardial infarction in the mouse. Proc. Natl. Acad. Sci. U. S. A. 108, 19725–19730 (2011).

4. Z. Huang, Z. Fu, W. Huang, K. Huang, Prognostic value of neutrophil-to-lymphocyte ratio in sepsis: A meta-analysis. W.B. Saunders [Preprint] (2020). 10.1016/j.ajem.2019.10.023.

5. S. Zhang, J. Diao, C. Qi, J. Jin, L. Li, X. Gao, L. Gong, W. Wu, Predictive value of neutrophil to lymphocyte ratio in patients with acute ST segment elevation myocardial infarction after percutaneous coronary intervention: A meta-analysis. BMC Cardiovasc. Disord. 18, 1–8 (2018).

6. B. H. Foy, T. M. Sundt, J. C. T. Carlson, A. D. Aguirre, J. M. Higgins, Human acute inflammatory recovery is defined by co-regulatory dynamics of white blood cell and platelet populations. Nat. Commun. 13, 1–10 (2022).

7. J. C. Marsh, D. R. Boggs, G. E. Cartwright, M. M. Wintrobe, Neutrophil kinetics in acute infection. J. Clin. Invest. 46, 1943–1953 (1967).

8. E. Rimmer, A. Garland, A. Kumar, S. Doucette, B. L. Houston, C. E. Menard, M. Leeies, A. F. Turgeon, S. Mahmud, D. S. Houston, R. Zarychanski, White blood cell count trajectory and mortality in septic shock: a historical cohort study. Canadian Journal of Anesthesia 69, 1230–1239 (2022).

9. B. Fernandes, Y. Hamaguchi, Automated Enumeration of Immature Granulocytes. Am. J. Clin. Pathol. 128, 454–463 (2007).

10. A. Hidalgo, E. R. Chilvers, C. Summers, L. Koenderman, The Neutrophil Life Cycle. Elsevier Ltd [Preprint] (2019). 10.1016/j.it.2019.04.013.

11. Y. Shi, S. Yang, M. Luo, W. D. Zhang, Z. P. Ke, Systematic analysis of coronary artery disease datasets revealed the potential biomarker and treatment target. Oncotarget 8, 54583–54591 (2017).

12. N. Zhang, X. Aiyasiding, W. jing Li,H. han Liao, Q. zhu Tang, Neutrophil degranulation and myocardial infarction. BioMed Central Ltd [Preprint] (2022). 10.1186/s12964-022-00824-4.

13. B. H. Foy, J. C. T. Carlson, A. D. Aguirre, J. M. Higgins, Platelet-white cell ratio is more strongly associated with mortality than other common risk ratios derived from complete blood counts. Nat. Commun. 16, 1113 (2025).

14. T. D. Miller, T. F. Christian, M. R. Hopfenspirger, D. O. Hodge, B. J. Gersh, R. J. Gibbons, Infarct size after acute myocardial infarction measured by quantitative tomographic 99mTc sestamibi imaging predicts subsequent mortality. Circulation 92, 334–341 (1995).

15. M. Licka, R. Zimmermann, J. Zehelein, T. J. Dengler, H. A. Katus, W. Kübler, Troponin T concentrations 72 hours after myocardial infarction as a serological estimate of infarct size. Heart 87, 520–524 (2002).

16. V. Roccaforte, G. Sabbatini, R. Panella, M. Daves, P. Formenti, M. Gotti, A. Galimberti, M. Spreafico, A. Piccin, G. Lippi, A. Pezzi, S. Pastori, The potential role of leukocytes cell population data (CPD) for diagnosing sepsis in adult patients admitted to the intensive care unit. Clin. Chem. Lab. Med. 63, 1031–1042 (2025).

17. S. H. Park, C.-J. Park, B.-R. Lee, K.-S. Nam, M.-J. Kim, M.-Y. Han, Y. J. Kim, Y.-U. Cho, S. Jang, Sepsis affects most routine and cell population data (CPD) obtained using the Sysmex XN-2000 blood cell analyzer: neutrophil-related CPD NE-SFL and NE-WY provide useful information for detecting sepsis. Int. J. Lab. Hematol. 37, 190–198 (2015).

18. N. Kumowski, S. Pabel, J. Grune, N. Momin, V. K. Ninh, L. Stengel, K. I. Mentkowski, Y. Iwamoto, Y. Zheng, I. H. Lee, J. Matthias, J. O. Wirth, F. E. Pulous, H. Seung, A. Paccalet, C. G. Muse, K. K. Y. Ting, P. Delgado, A. J. M. Lewis, V. Kaushal, A. Kreso, D. Brown, S. Hayat, R. Kramann, F. K. Swirski, K. Naxerova, D. C. Propheter, L. V. Hooper, M. A. Moskowitz, K. R. King, N. Rosenthal, M. Hulsmans, M. Nahrendorf, Resistin-like molecule γ attacks cardiomyocyte membranes and promotes ventricular tachycardia. Science (1979). 389, 1043–1048 (2025).

19. A. Tavosanis, Restraining emergency hematopoiesis preserves cardiac function after infarction. Nature Cardiovascular Research 2025 4:6 4, 650–650 (2025).

20. S. Zhang, A. Paccalet, D. Rohde, S. Cremer, M. Hulsmans, I. H. Lee, K. Mentkowski, J. Grune, M. J. Schloss, L. Honold, Y. Iwamoto, Y. Zheng, M. A. Bredella, C. Buckless, B. Ghoshhajra, V. Thondapu, A. M. van der Laan, J. J. Piek, H. W. M. Niessen, F. Pallante, R. Carnevale, S. Perrotta, D. Carnevale, O. Iborra-Egea, C. Muñoz-Guijosa, C. Galvez-Monton, A. Bayes-Genis, C. Vidoudez, S. A. Trauger, D. T. Scadden, F. K. Swirski, M. A. Moskowitz, K. Naxerova, M. Nahrendorf, Bone marrow adipocytes fuel emergency hematopoiesis after myocardial infarction. Nature Cardiovascular Research 2023 2:12 2, 1277–1290 (2023).

21. S. L. Puhl, S. Steffens, Neutrophils in Post-myocardial Infarction Inflammation: Damage vs. Resolution? Frontiers Media S.A. [Preprint] (2019). 10.3389/fcvm.2019.00025.

22. L. A. Heger, N. Schommer, S. Van Bruggen, C. E. Sheehy, W. Chan, D. D. Wagner, Neutrophil NLRP3 promotes cardiac injury following acute myocardial infarction through IL-1β production, VWF release and NET deposition in the myocardium. Sci. Rep. 14, 14524 (2024).

23. C. Piot, P. Croisille, P. Staat, H. Thibault, G. Rioufol, N. Mewton, R. Elbelghiti, T. T. Cung, E. Bonnefoy, D. Angoulvant, C. Macia, F. Raczka, C. Sportouch, G. Gahide, G. Finet, X. André-Fouët, D. Revel, G. Kirkorian, J.-P. Monassier, G. Derumeaux, M. Ovize, Effect of Cyclosporine on Reperfusion Injury in Acute Myocardial Infarction. New England Journal of Medicine 359, 473–481 (2008).

24. N. Mewton, F. Roubille, D. Bresson, C. Prieur, C. Bouleti, T. Bochaton, F. Ivanes, O. Dubreuil, L. Biere, A. Hayek, F. Derimay, M. Akodad, B. Alos, L. Haider, N. El Jonhy, R. Daw, C. De Bourguignon, C. Dhelens, G. Finet, E. Bonnefoy-Cudraz, G. Bidaux, F. Boutitie, D. Maucort-Boulch, P. Croisille, G. Rioufol, F. Prunier, D. Angoulvant, “Effect of Colchicine on Myocardial Injury in Acute Myocardial Infarction” in Circulation (Lippincott Williams and Wilkins, 2021; https://www.ahajournals.org/doi/suppl/10.1161/CIRCULATIONAHA.121.056177.) vol. 144, pp. 859–869.

25. M. Panahi, A. Papanikolaou, A. Torabi, J. G. Zhang, H. Khan, A. Vazir, M. G. Hasham, J. G. F. Cleland, N. A. Rosenthal, S. E. Harding, S. Sattler, Immunomodulatory interventions in myocardial infarction and heart failure: A systematic review of clinical trials and meta-analysis of IL-1 inhibition. Oxford University Press [Preprint] (2018). 10.1093/cvr/cvy145.

26. P. M. Ridker, B. M. Everett, T. Thuren, J. G. MacFadyen, W. H. Chang, C. Ballantyne, F. Fonseca, J. Nicolau, W. Koenig, S. D. Anker, J. J. P. Kastelein, J. H. Cornel, P. Pais, D. Pella, J. Genest, R. Cifkova, A. Lorenzatti, T. Forster, Z. Kobalava, L. Vida-Simiti, M. Flather, H. Shimokawa, H. Ogawa, M. Dellborg, P. R. F. Rossi, R. P. T. Troquay, P. Libby, R. J. Glynn, Antiinflammatory Therapy with Canakinumab for Atherosclerotic Disease. New England Journal of Medicine 377, 1119–1131 (2017).

27. A. Abbate, C. R. Trankle, L. F. Buckley, M. J. Lipinski, D. Appleton, D. Kadariya, J. M. Canada, S. Carbone, C. S. Roberts, N. Abouzaki, R. Melchior, S. Christopher, J. Turlington, G. Mueller, J. Garnett, C. Thomas, R. Markley, G. F. Wohlford, L. Puckett, H. M. de Chazal, J. G. Chiabrando, E. Bressi, M. G. Del Buono, A. Schatz, C. Vo, D. L. Dixon, G. G. Biondi-Zoccai, M. C. Kontos, B. W. Van Tassell, Interleukin-1 blockade inhibits the acute inflammatory response in patients with st-segment–elevation myocardial infarction. J. Am. Heart Assoc. 9 (2020).

28. J. Rettkowski, M. C. Romero-Mulero, I. Singh, C. Wadle, J. Wrobel, D. Chiang, N. Hoppe, J. Mess, K. Schönberger, M. E. Lalioti, K. Jäcklein, B. SilvaRego, T. Bühler, N. Karabacz, M. Egg, H. Demollin, N. Obier, Y. W. Zhang, C. Jülicher, A. Hetkamp, M. Czerny, M. J. Jones, H. Seung, R. Jain, C. von zur Mühlen, A. Maier, A. Lother, I. Hilgendorf, P. van Galen, A. Kreso, D. Westermann, A. E. Rodriguez-Fraticelli, T. Heidt, N. Cabezas-Wallscheid, Modulation of bone marrow haematopoietic stem cell activity as a therapeutic strategy after myocardial infarction: a preclinical study. Nature Cell Biology 2025 27:4 27, 591–604 (2025).

29. B. H. Foy, R. Petherbridge, M. T. Roth, C. Zhang, D. C. De Souza, C. Mow, H. R. Patel, C. H. Patel, S. N. Ho, E. Lam, C. E. Powe, R. P. Hasserjian, K. J. Karczewski, V. Tozzo, J. M. Higgins, Haematological setpoints are a stable and patient-specific deep phenotype. Nature 637, 430–438 (2025).

