## Supplemental Material for "WBC population dynamics differ in response to acute ischemic and infectious insults and can discriminate clinically maladaptive responses to myocardial infarction"

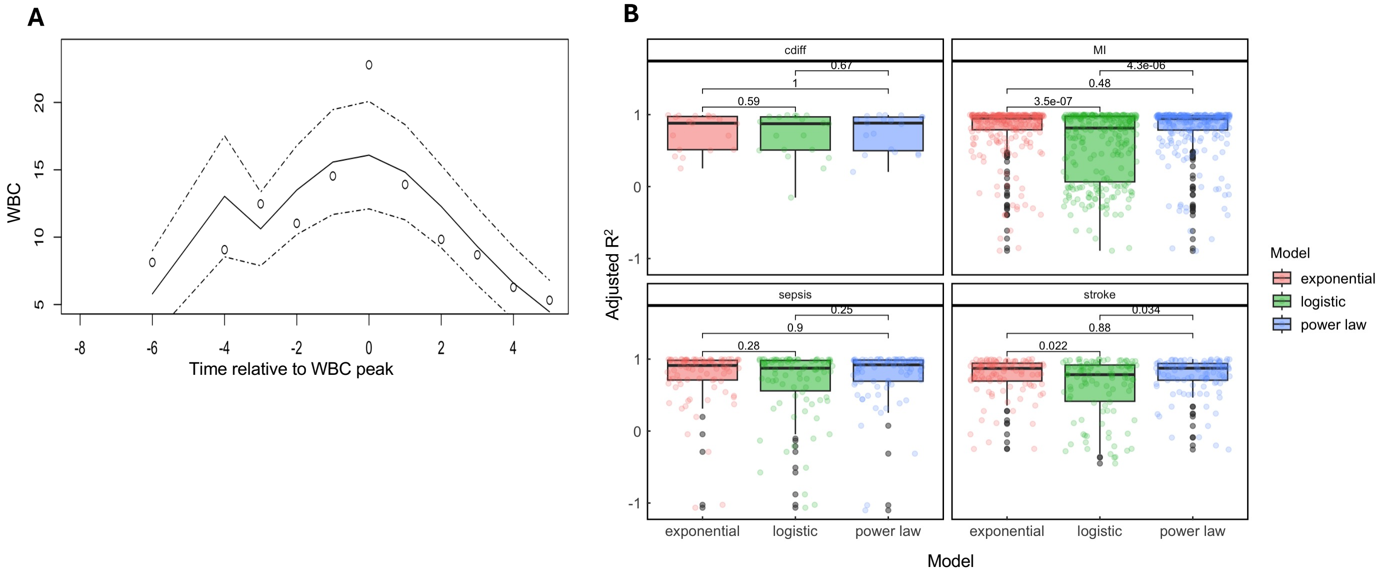


**Fig. S1. Models of leukocyte response phase. A.** Representative patient WBC values (points) around the time of WBC peak ($t = 0$), with superimposed fit for an exponential growth model with a carrying capacity (Supplemental material, Section S1). Solid line gives the median fit, while dashed lines give the 95% credible interval by Markov Chain Monte Carlo. **B.** Etiology-specific comparison of exponential fit with two additional statistical models: logistic growth and power law dynamics. P-values determined by Wilcoxon rank-sum test.

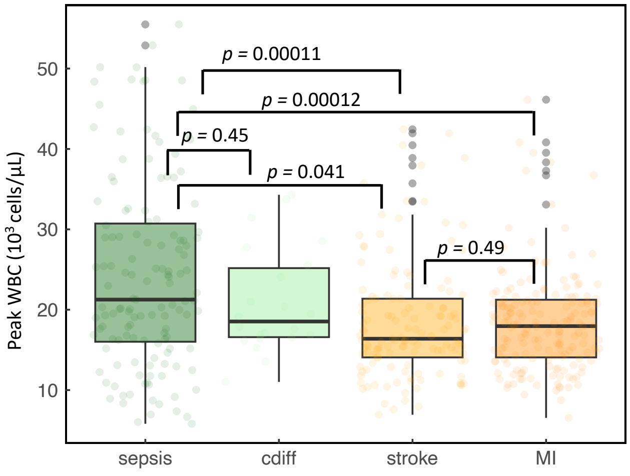

**Fig. S2. Peak absolute WBC by etiology.** Boxplots showing distribution of peak WBC in each etiology-sepcific patient cohort. The black horizontal line denotes the median, and the shaded area gives the IQR. *P* values for pairwise comparisons of etiologies calculated via Wilcoxon rank sum test, $\alpha = 0.05$.

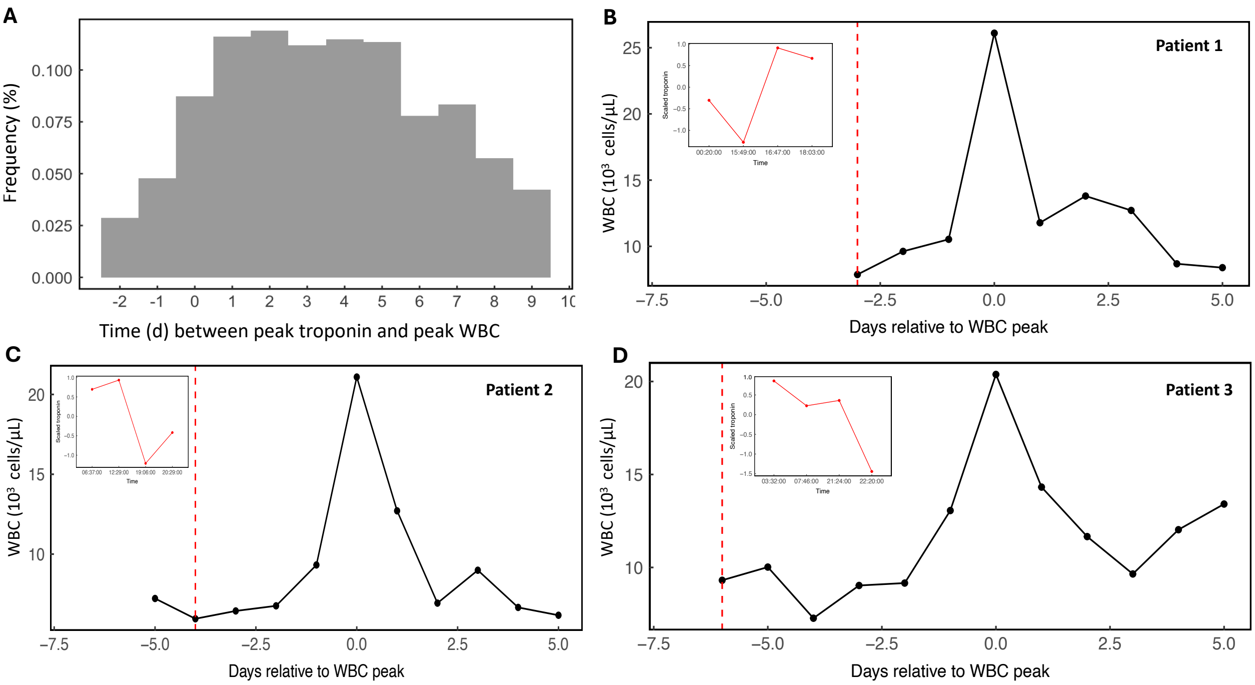


**Fig. S3. Relationship between early leukocyte response dynamics and timing of ischemic event for patients with AMI. A.** Distribution of time (days) between peak troponin and time of WBC peak (median = 3d) for $n$ = 910 patients in the AMI cohort for whom troponins were available in the EMR during admission (for 196 AMI patients that met inclusion criteria, hs-troponin was not available with routine CBC data, for example if collected at an outside emergency department prior to transfer for admission to this tertiary care center). **B-D.** Representative individual WBC trajectories for patients in the AMI cohort, with vertical dashed line giving the date of peak troponin. Inset plots show troponin dynamics on the date of troponin peak.

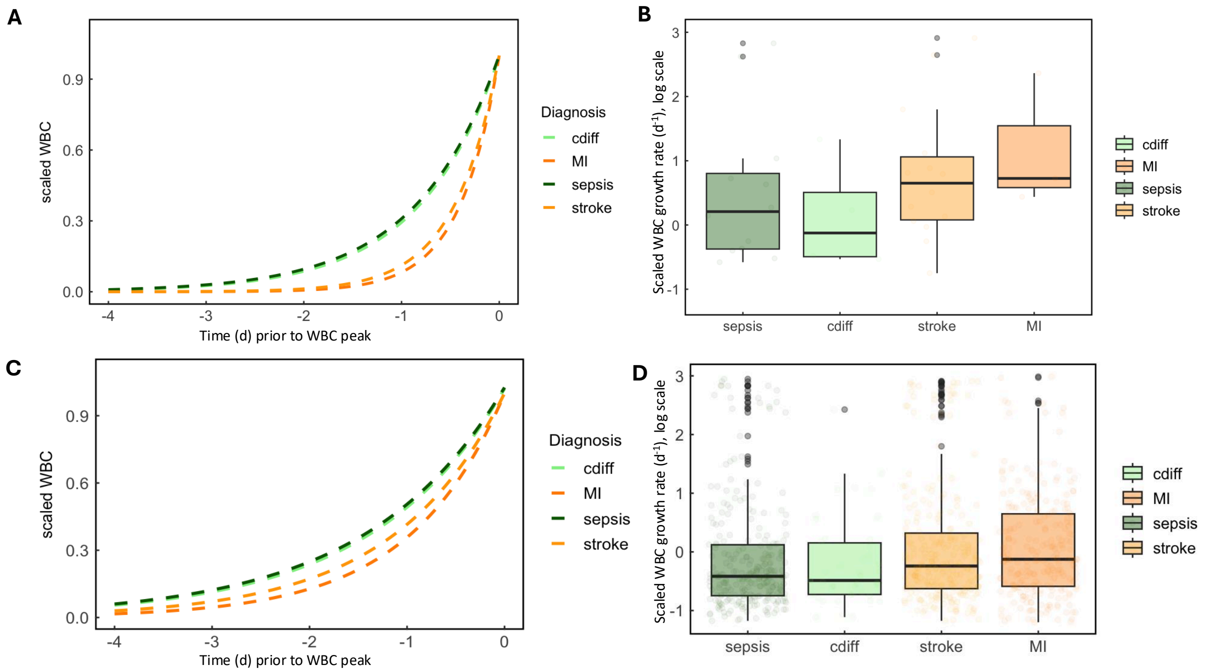


**Fig. S4. Alternate classifications of the "favorable trajectory" cohort.** Inferred leukocyte response-phase dynamics using goodness-of-fit (adjusted $R^2$) thresholds of 0.95 (**A** and **B**, conservative definition) and 0.7 (**C** and **D**, relaxed definition), respectively. **A**. Exponential fit of etiology-specific WBC response trajectories across individuals for the conservative adjusted $R^2$ threshold. **B**. Boxplot of inferred exponential WBC decay rates for individuals by etiology under the conservative adjusted $R^2$ threshold. **C**. Exponential fit of etiology-specific WBC response trajectories across individuals for the relaxed adjusted $R^2$ threshold. **D**. Boxplot of inferred exponential WBC decay rates for individuals by etiology under the relaxed adjusted $R^2$ threshold.

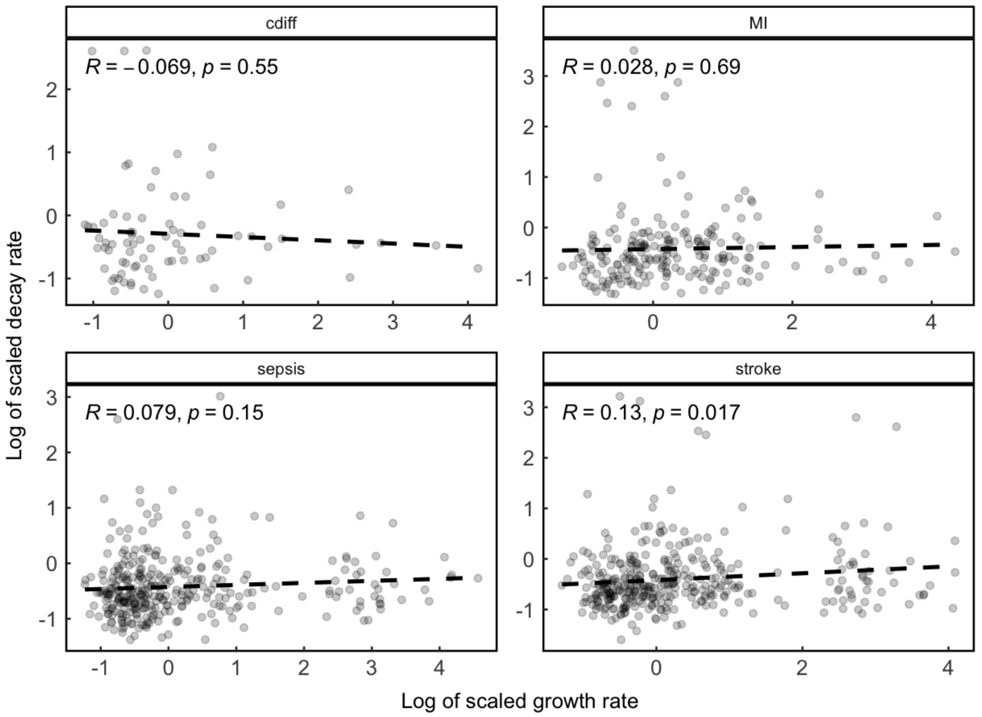


**Fig. S5. Leukocyte growth is independent of leukocyte decay.** Inferred scaled leukocyte decay rate (units are 1/day on a log scale) vs. inferred leukocyte response rate (units are 1/day on a log scale) across individuals for each etiology in the 'favorable trajectory' cohort (Methods).

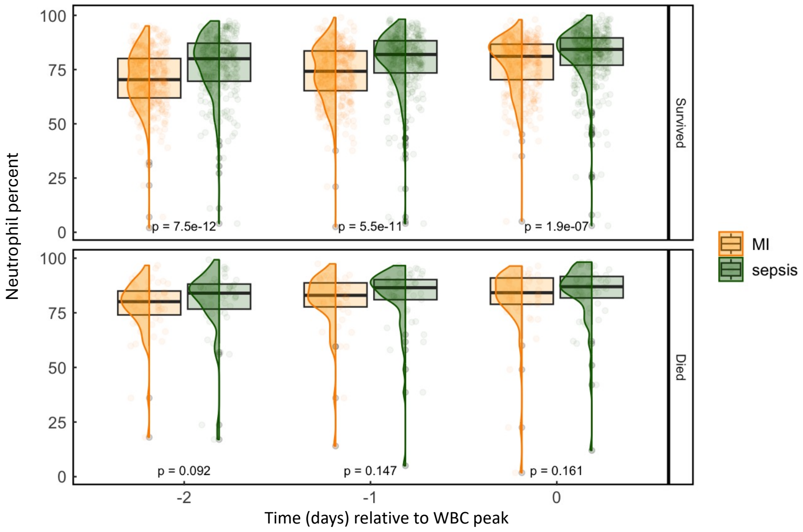


**Fig. S6. Comparison of neutrophil fraction between AMI and sepsis cohorts close to WBC peak, stratified by short-term mortality.** Boxplots (bolded line, median; box limits, IQR) showing %neutrophil among surviving patients (top panel) and non-surivors (bottom panel) over the 3 days leading to WBC peak, stratified by etiology. Points denote individual values.

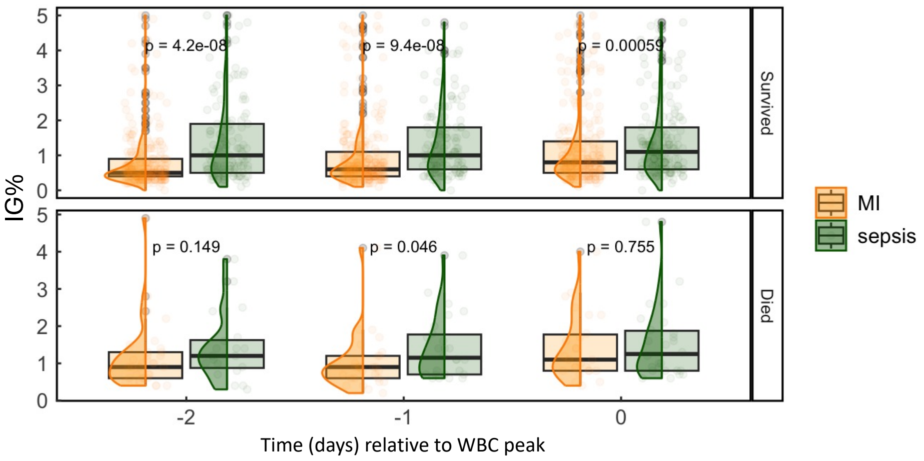


**Fig. S7. Comparison of immature granulocyte fraction between AMI and sepsis cohorts close to WBC peak, stratified by short-term mortality.** Boxplots (bolded line, median; box limits, IQR) showing %neutrophil among surviving patients (top panel) and non-surivors (bottom panel) over the 3 days leading to WBC peak, stratified by etiology. Points denote individual values.

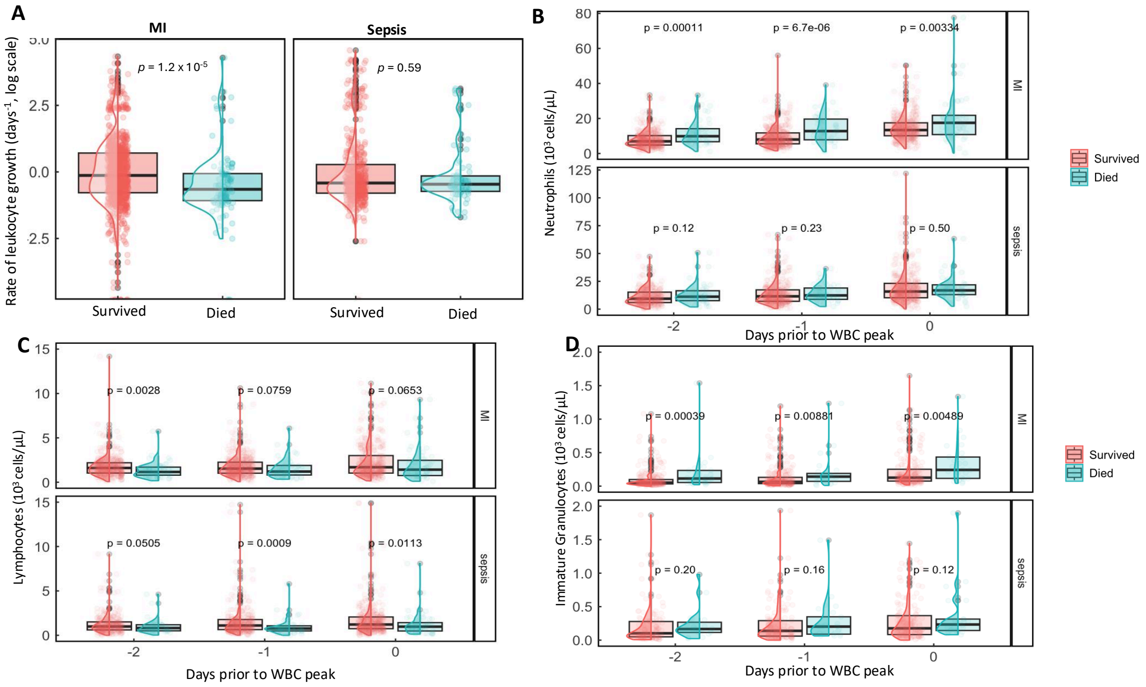


**Fig. S8. A.** Boxplots and overlaid distributions showing exponential WBC growth rates in AMI (left) and sepsis (right), stratified according to short-term mortality. Black horizontal lines give the median across patients, and shaded boxes give 95% confidence intervals. *P*-values calculated by Wilcoxon rank sum test (alpha=0.05). Outliers are shown as dots, and distributions are overlaid on the boxplots. **B.** Boxplots and overlaid distributions showing absolute neutrophil counts among AMI patients (top) and sepsis patients (bottom) stratified by short-term mortality on each of the three days prior to WBC peak. **C.** Boxplots and overlaid distributions showing absolute lymphocyte counts among AMI patients (top) and sepsis patients (bottom) stratified by short-term mortality on each of the three days prior to WBC peak. **D.**

Boxplots and overlaid distributions showing absolute immature granulocyte (IG) counts among AMI patients (top) and sepsis patients (bottom) stratified by short-term mortality on each of the three days prior to WBC peak.

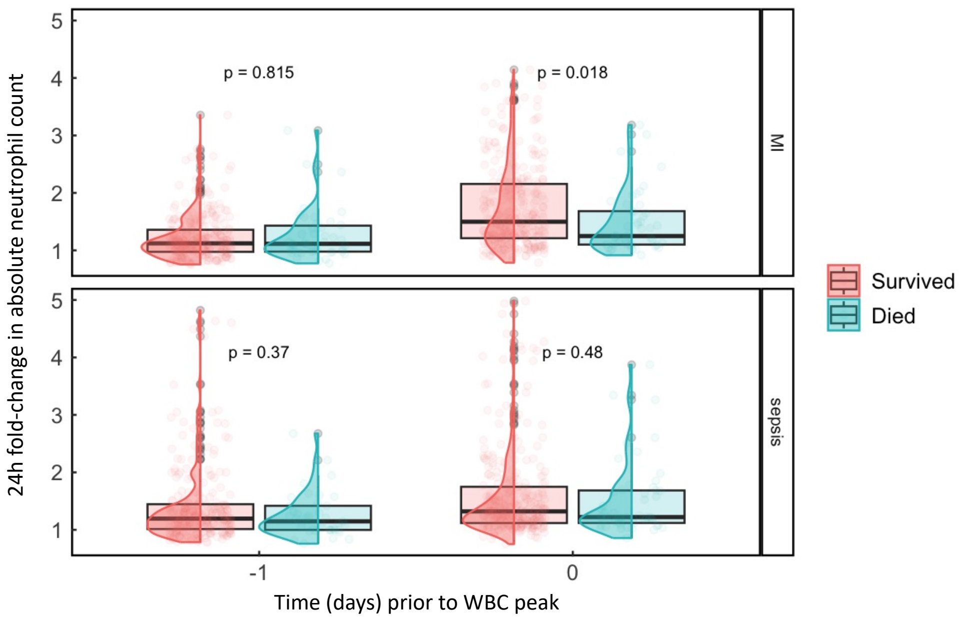


**Fig. S9. Neutrophil acceleration close to WBC peak among patients with AMI and sepsis, stratified by short-term mortality.** Boxplots (bolded line, median; box limits, IQR) showing 24h fold-change in absolute neutrophil count among patients with AMI (top panel) and sepsis (bottom panel) over the day prior to WBC peak and the day of WBC peak. Points denote individual values.

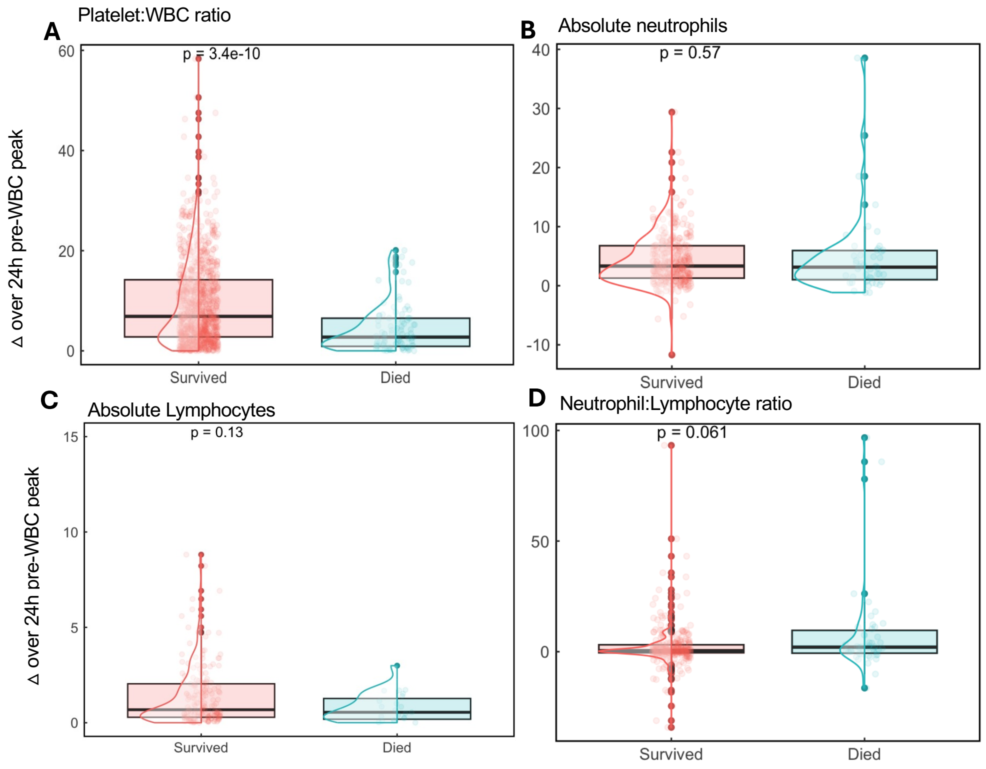


**Fig. S10 Change (Δ) in hematologic markers around WBC peak among patients with myocardial infarction. A**. Boxplot (bolded line, median; box limits, IQR) showing change in platelet:WBC ratio (PWR) across individuals, stratified by short-term mortality. **B**. Change in absolute neutrophil count stratified by short-term mortality. **C**. Change in absolute lymphocyte count stratified by short-term mortality. **D**. Change in neutrophil:lymphocyte ratio (NLR) stratified by short-term mortality.

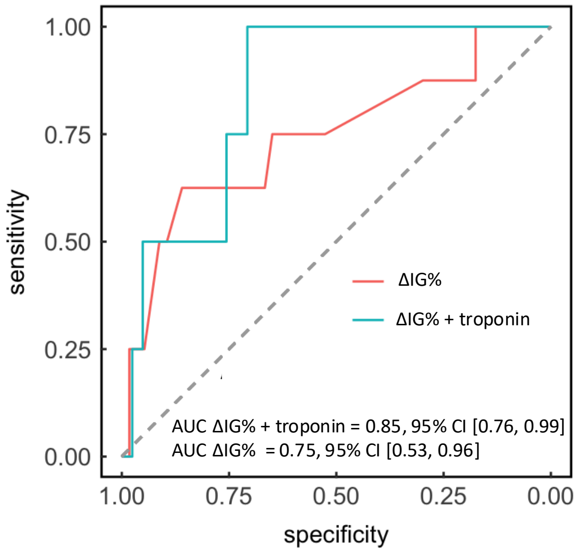


**Fig. S11.** Receiver operating characteristic (ROC) curves showing performance of $\Delta_{IG}$ alone as a predictor of 30d mortality compared to performance of a model that includes both $\Delta_{IG}$ and scaled troponin. Area under the curve (AUC) constructed by bootstrapping (10,000 replicates).

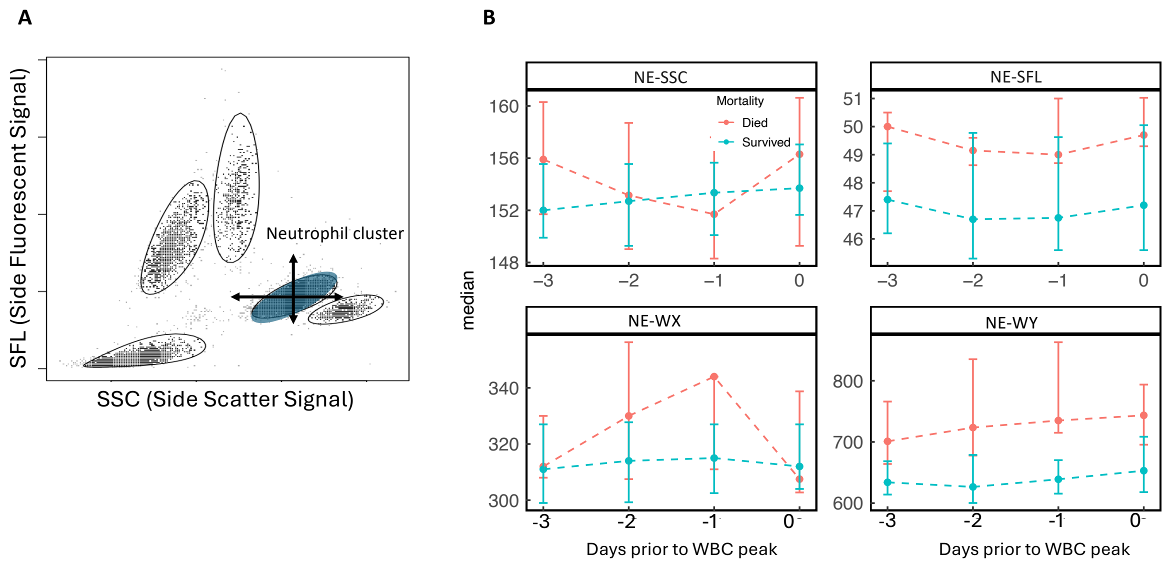


**Fig. S12. Analysis of single cell distribution parameters among patients with AMI close to WBC peak. A.** Representative single cell scatter of leukocyte populations from one individual at one time point. Blue shaded area denotes the neutrophil cluster. **B.** Median (point) and IQR (error bars) across AMI patients of single-cell parameters along the x- axis (neutrophil single cell scatter, NE-SSC, and neutrophil side scatter dispersion, NE-WX) and y-axis (neutrophil fluorscence intensity, NE-SFL, and neutrophil fluorescence dispersion, NE-WY) of the leukocyte scattergram. Results stratified according to 30d mortality.

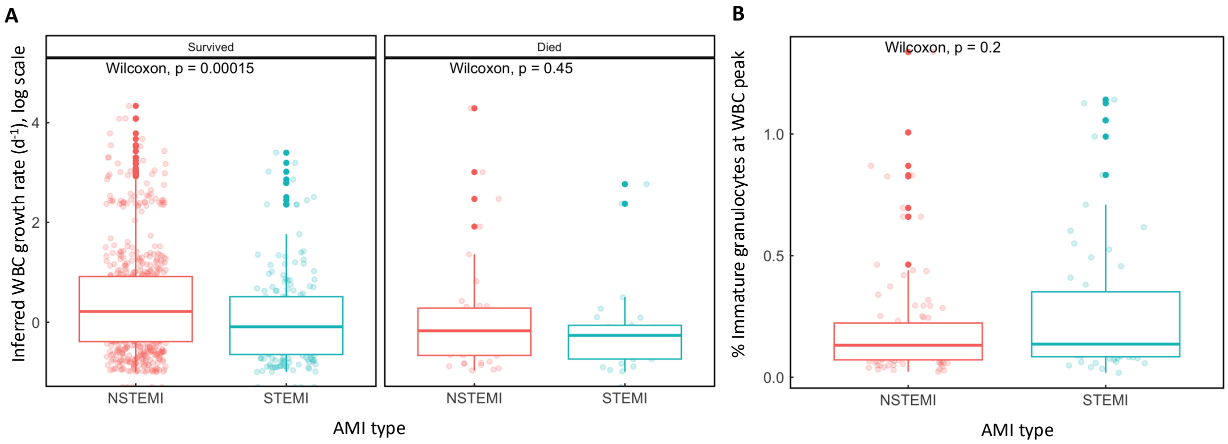


**Fig. S13. Response-phase leukocyte dynamics according to AMI type. A**. Boxplots of inferred leukocyte response rate among patients with acute myocardial infarction according to AMI type (NSTEMI vs. STEMI), stratified by short-term mortality. **B**. Boxplot of % Immature granulocytes (of total WBC) at the time of WBC peak, according to AMI type (NSTEMI vs. STEMI). Points give individual values.

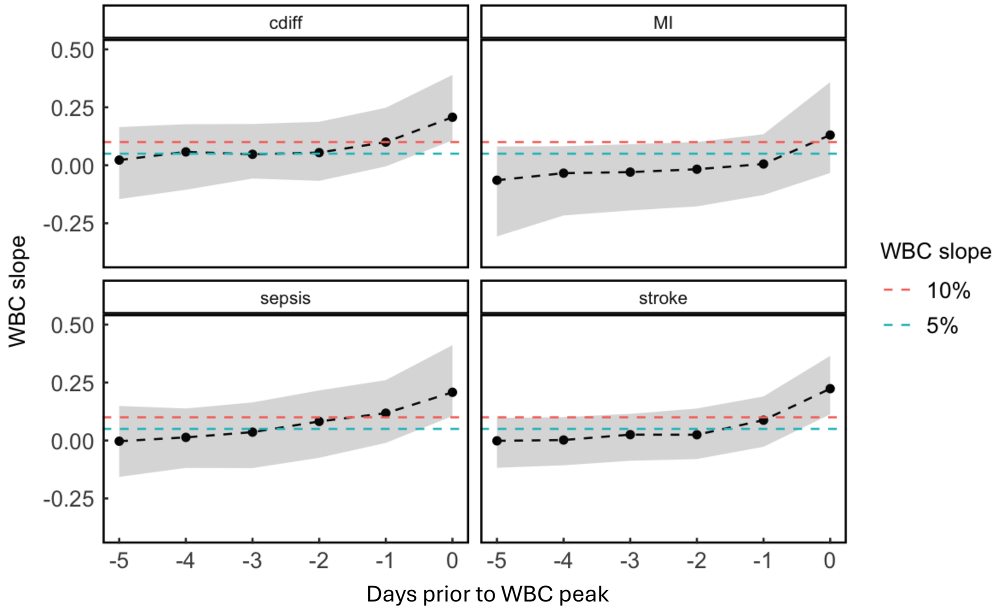


**Fig. S14. Determining the start of the WBC response phase.** Slope of WBC growth (ΔWBC/Δt, units $10^3$ cells/µl/day) over five days prior to WBC peak**.** Black dashed lines give the population median, and gray shaded area denotes the interquartile range. Red and blue dashed lines correspond to 5% and 10% positive increases in slope, respectively.

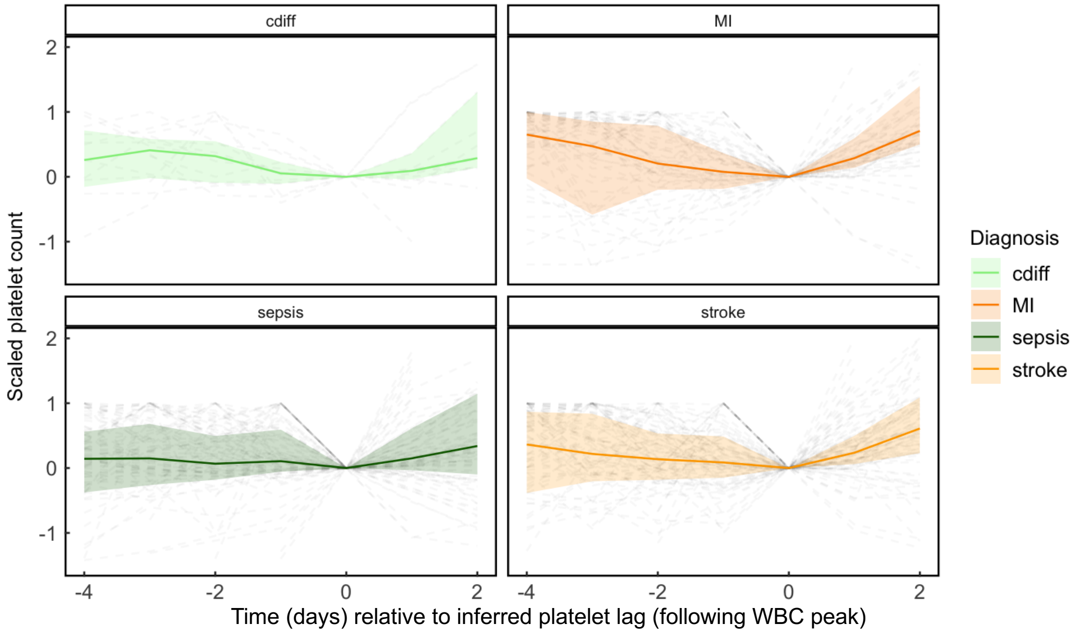


**Fig. S15. Heterogeneous etiology-specific platelet trajectories.** Solid line gives medians across individuals at each timepoint. Shaded area represents the inter-quartile range at each timepoint, and light grey dotted lines show individual trajectories. Platelets were scaled and shifted around the inferred platelet lag (t = 0 days) as specified in the Methods.

**Section S1. Functional form of response-phase leukocyte growth.**
Response-phase leukocyte dynamics were empirically well-represented by exponential growth curves among patients that met inclusion criteria (main text, Fig. 1D, inset). We compared the exponential model against several other models of the leukocyte response phase. First, we tried a model for exponential growth with a carrying capacity

$$\frac{dWBC}{dt} = r * WBC\left(1 - \frac{WBC}{K}\right),$$

modeling WBC population growth with initial rate *r* and carrying capacity *K.* This model was fitted to observed individual WBC trajectories via Markov Chain Monte Carlo, with a negative binomial observation process. Starting parameters for the fitting routine were chosen for each individual from the exponential model fits (rate *r*) and the maximum observed WBC count (capacity *K)*. The carrying capacity model consistently under-estimated the peak WBC (e.g. Fig. S1A), failing to capture the key acceleration of the leukocyte response phase. We next compared two additional empirical statistical models – power law dynamics and logistic growth - against the exponential fit. For each etiology, the exponential growth model provided superior or non-inferior fits for the leukocyte response phase compared to the other models (Fig. S1B).

**Section S2. Defining a favorable leukocyte trajectory.** To investigate baseline differences in leukocyte dynamics following acute infection and ischemia, we first defined a cohort of patients that exhibited orderly, or “favorable” dynamics. Building on our prior analysis of the WBC recovery phase in this cohort[6], we focused on patients who had well-fitting WBC and PLT recovery dynamics (main text, Methods), and well-fitting exponential growth during the WBC response phase, using a goodness of fit threshold of adjusted $R^2 > 0.85$. We assessed the robustness of these observed etiology-specific trends to alternative definitions of the “favorable trajectory cohort” using alternative goodness-of-fit thresholds:

1. Conservative definition (adjusted $R^2 > 0.95$ for exponential leukocyte response-phase growth, Fig. S5, A and B).
2. Relaxed definition (adjusted $R^2 > 0.7$ for exponential leukocyte response-phase growth, Fig. S5, C and D).

Results were qualitatively similar to the etiology-specific trends observed in the main analysis (main text, Fig. 1).

**Section S3. Example clinical courses for patients with “favorable” and “unfavorable” leukocyte trajectories.**
As above, patients in the “favorable trajectory cohort” were defined according to short-term (30d) mortality and features of the leukocyte response (Methods, Section S2). Here, to provide clinical context, we contrast the clinical course of an AMI patient identified within the favorable trajectory cohort (patient X) with that of an AMI patient who did not meet criteria for the favorable trajectory cohort (Patient Y) and did not survive to day 30. Patient X was a 61 year old male with a history of hypertension, insulin dependent diabetes, prior venous thromboembolism, and multivessel coronary artery disease admitted with anterior wall NSTEMI. The patient remained hemodynamically stable on presentation and was admitted for medical management. The patient underwent left heart catheterization on hospital day 2, with successful balloon angioplasty and stenting of the obtuse marginal artery 2, and recovered uneventfully post-procedurally. A post-procedural trans-thoracic echocardiogram showed preserved left ventricular ejection fraction compared to pre-admission baseline. Patient Y was a 71 year old male with a past medical history of multivessel CAD and chronic obstructive lung disease admitted with chest pain and

anterior ST segment elevation myocardial infarction. Cardiac catheterization revealed distal left anterior descending artery occlusion, which was stented. Several days post-procedurally, the patient experienced ventricular arrhythmia (monomorphic ventricular tachycardia) and subsequent cardiac decompensation, with new left ventricular dysfunction, shock, and acute respiratory failure, eventually progressing to multiorgan failure and death during the admission.

**Section S4. Impact of type of myocardial infarction on leukocyte activity close to WBC peak.**
Of $n = 1106$ patients with myocardial infarction included in the analysis, 267 (26.0%) were admitted with a diagnosis of ST-segment elevation MI (STEMI), while 756 (74%) were admitted with a diagnosis of Type II (demand ischemia) NSTEMI. The predominance of NSTEMI captured in the analysis likely reflects the observation structure necessary to model response-phase leukocyte dynamics (observed WBC counts over 3d prior to WBC peak). Many patients with STEMI present with maximum WBC count on the day of acute vascular occlusion, with sharp and sustained WBC decrease following percutaneous or surgical intervention. Accordingly, among surviving patients, the inferred rate of WBC growth during the leukocyte response phase was significantly higher among patients with STEMI compared to NSTEMI (Fig. S10A). However, among patients who did not survive, the rate of leukocyte growth was slower and similar regardless of AMI type. Also, there was no significant difference in immature granulocyte activity close to WBC peak (Fig. S10B).

**Section S5. Changes in hematologic cell populations close to WBC peak and impact on short-term mortality risk.** Changes in the immature granulocyte population (ΔIGF, main text) over the 24h prior to WBC peak were strongly associated with short-term mortality in patients with AMI (main text). The survival classification according to ΔIGF was stronger than that according to analogous changes in other leukocyte populations and markers (Fig. S5), including neutrophil:lymphocyte ratio. Consistent with a recent complementary investigation,[13] change in the platelet:WBC ratio (PWR) over 24h prior to WBC peak was also associated with short-term mortality in our AMI population (Fig. S5A). In a generalized linear model, ΔIG showed similar predictive association for short-term mortality (OR 2.04, 95% CI [1.41, 3.13]) as ΔPWR (OR 0.37, 95% CI [0.25, 0.52]).
